# Associations of Smoking Status and Leisure-time Physical Activity with Waist Circumference Change – Ten-year Follow-up among Twin Adults

**DOI:** 10.1101/2024.09.26.24314418

**Authors:** Piirtola Maarit, Filippone Eeva-Liisa, Ranjit Anu, Kinnunen Taru, Kaprio Jaakko, Korhonen Tellervo

## Abstract

**BACKGROUND:** This cohort study investigated the associations of smoking status and leisure-time physical activity (LTPA) with weight circumference (WC) change.

**METHODS:** In the FinnTwin16 cohort, 3,322 twins (46% men) reported smoking status, LTPA, and WC in early adulthood and 10 years later providing information on essential covariates at baseline. The effects of smoking and LTPA (metabolic equivalent tasks [MET]-h/week) on WC change (cm) were estimated by modelling WC value at the end of follow-up and adjusted for baseline WC in linear regression models. Within-pair associations were analyzed using linear fixed-effect regressions among 660 dizygotic and 390 monozygotic twin pairs.

**RESULTS:** During the 10-year follow-up, 36.4% (n=273) of baseline daily smokers quit smoking. Among those who quit daily smoking, the mean WC increase was 8.4 cm (SD 8.1). Quitters who smoked daily at baseline increased WC by about 2 cm more than continuing smokers (adjusted β 2.04; 95% CI 0.94, 3.14). This association was not robust after shared familial influences were controlled for. In general, the participants decreased LTPA during follow-up, except the quitters with the mean LTPA increase of 5.0 MET-h/week (SD 35.0). Independently of smoking status, each additional MET-h/week was associated with 0.06 cm less WC increase (adjusted β -0.06; 95% CI - 0.07, -0.05). This association was replicated in the within-pair analyses.

**CONCLUSIONS:** Smoking cessation seems to be associated with WC increase, but familial confounding may be involved in this process. LTPA appears to mitigate increases in WC independently of smoking status and familial influences.

## INTRODUCTION

Although smoking cessation has numerous health benefits, individuals quitting smoking tend to gain weight^1–3^. On average, the first post-cessation year weight gain is 3-6 kilograms, but 10-15% experience more weight gain^3^ ^4^. This is modified by age, sex, pre-cessation Body Mass Index (BMI) and heavy smoking^5^ ^6^. Negative consequences of weight gain main reduce the benefits of smoking cessation^7^ ^8^.

Post-cessation weight changes are well documented. However, weight may not be the most informative health-related measurement. Measures of abdominal obesity, such as waist circumference (WC), may be a better predictors of weight-related health conditions than weight^9,10^. There is short-term evidence of increased abdominal obesity after smoking cessation^11–13^. However, verification on long-term post-cessation changes in abdominal obesity is limited.

Leisure-time physical activity (LTPA) has been shown to inhibit weight gain in general^14^. Evidence of whether LTPA could impact post-cessation weight gain has been inconsistent^15^. It has been suggested that exercise may prevent weight gain in the long run^15^. Less is known about the impact of LTPA on post-cessation change in abdominal obesity. However, in a smoking cessation trial including exercise intervention, abstinence was followed by moderate weight gain, but not by increase in relation between visceral and abdominal fat^16^.

Finally, familial influences, including shared genetic and environmental factors, may confound the associations of smoking behavior and physical activity with metabolic reactions of the body^17–19^. In epidemiological research, twin samples are therefore valuable in testing the role of familial factors^20^.

## AIMS

Our aims were to investigate how smoking behavior, especially smoking cessation, is associated with WC change during the 10-year follow-up and whether higher amount of LTPA is associated with lower level of WC increase. Our further aim was to test if the associations are independent of familial influences.

## METHODS

### Sample

The data is based on the Finnish population sample, FinnTwin16 cohort^21^. We used two surveys conducted in 2000-2003 (baseline) and 2010-2012 (follow-up) (Figure 1a). At baseline 5 240 (46% men, mean age 24 years, born in 1975-79), and at follow-up, 4,397 (45% men, mean age 34 years) twins completed the surveys. Data were collected by questionnaires, including self-measurements of WC. Analyses are based on 3,322 twins (46% men) who reported their smoking status, LTPA, and WC in both surveys and provided information on essential covariates at baseline. Characteristics of the participants by long-term smoking status are presented in Supplement Tables 1 and 2.

**Figure 1a.**
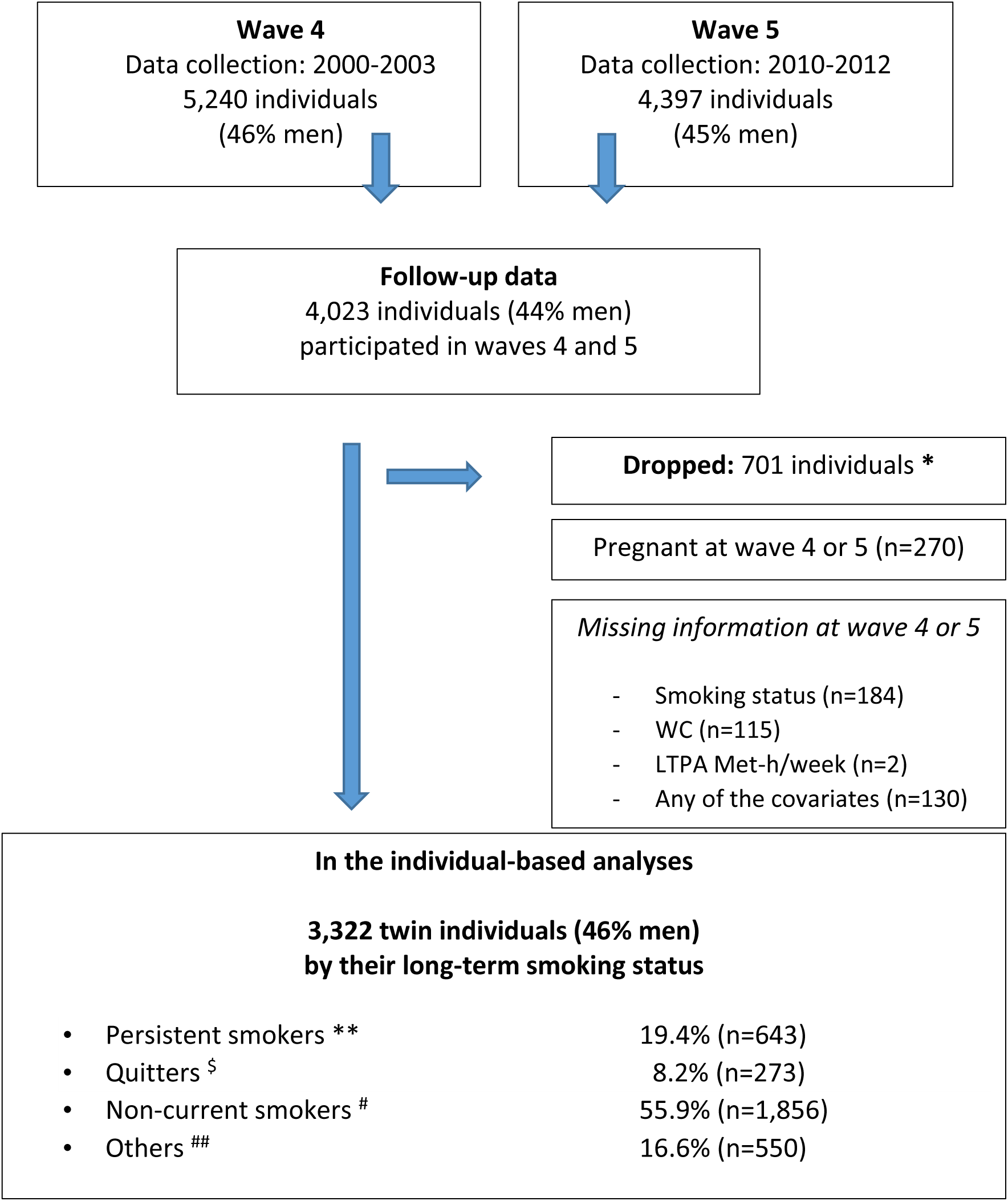
The flow chart of individuals included in the individual-based analyses by their long-term smoking status. WC= waist circumference, LTPA= leisure time physical activity, Met-h/week=metabolic equivalent hours/week *Dropped in listed order. ** Persistent daily or occasional smoking. ^$^ Quitting from daily smoking. ^#^ Quitting from occasional smoking, consistent former smoking, never smoking. ^##^ Miscellaneous smoking statuses (initiators, reducers, relapses, increasers, and other changes in smoking status during wave 4 and wave 5).

Data includes 1,085 full twin pairs who fulfilled the inclusion criteria and both siblings participated both data collections. Of those twin pairs, 390 were monozygotic (MZ), 660 were dizygotic (DZ) (328 same-sex (SS), 332 opposite-sex (OS)), and 35 SS pairs with uncertain zygosity (Figure 1b). For the within-pair analyses, we used data from the 1,050 pairs with known zygosity. The determination of zygosity was done using a validated questionnaire method^22^ supplemented with genetic marker information for some twin pairs.

**Figure 1b:**
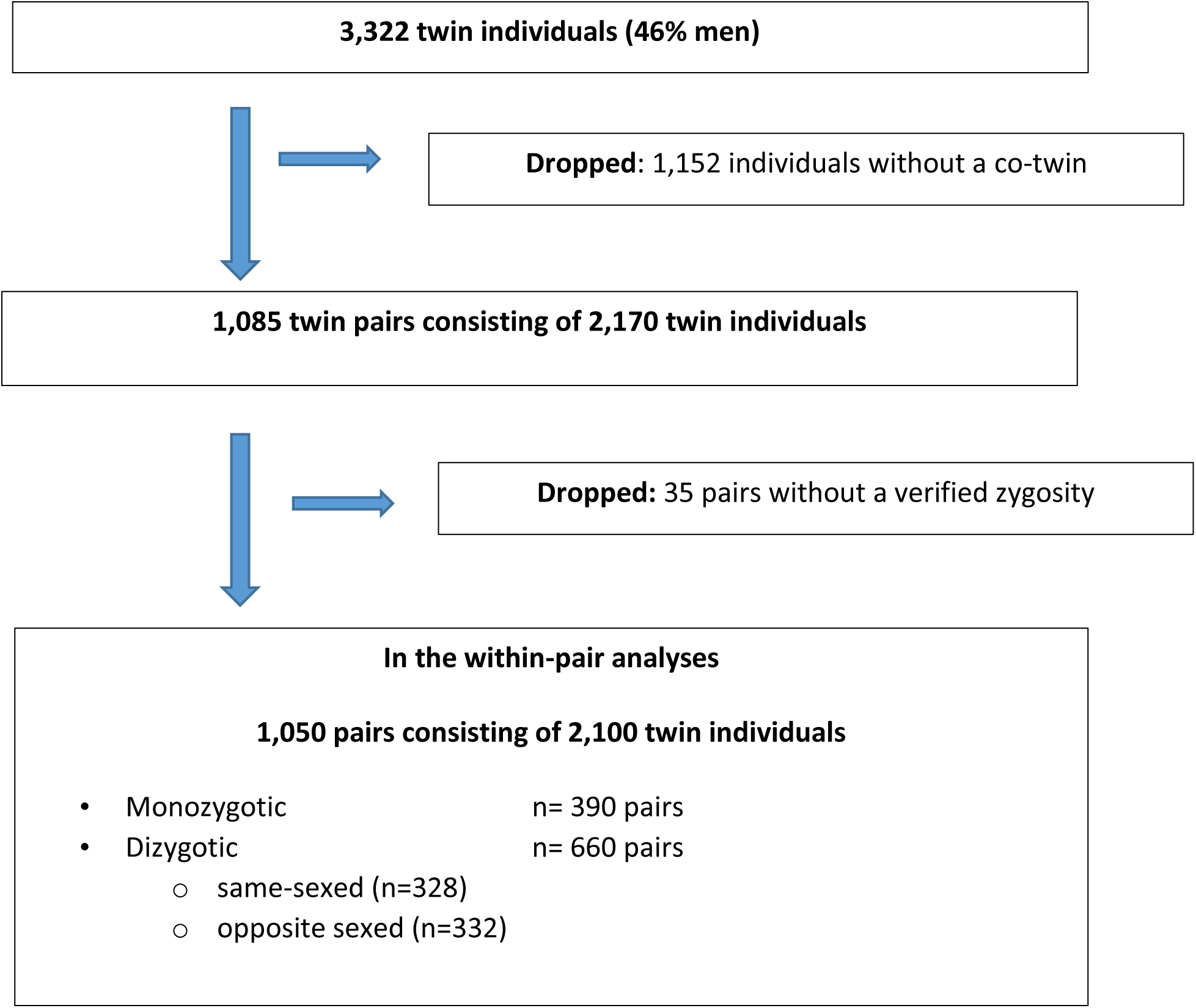
The flow chart of the inclusion of twin individuals into the within-pair analyses.

### Dependent variable

Abdominal obesity was operationalized with a self-measured WC in centimeters at baseline and follow-up. Participants received pictorial instruction and a tape measure^23^. Pictorial instruction is shown in Supplement Figure 1. Self-measured WC shows reasonable accuracy in Finnish data^23^. For descriptive purpose, change in WC was calculated as the difference in centimeters between the follow-up and baseline values. In the regression analyses, the dependent variable was the WC value at the end of follow-up while the baseline WC was adjusted for, and the result can be interpreted to reflect change in WC.

### Independent variables

#### Smoking status

At baseline, smoking status was asked with a categorical question: smoking daily (≥20, 10-19, ≤9 cigarettes), smoking occasionally (at least once week, less frequently than each week), has quit smoking, being a never-smoker. At follow-up, smoking status question included: smoking daily, smoking weekly, smoking less often than weekly, has quit smoking, never smoked. The participants were then categorized into the four long-term smoking status groups. Persons who were persistently smoking daily or occasionally were classified as ’Persistent smokers’ (n=643/19.3%), those who were daily smokers at baseline but quit during the follow-up as ’Quitters’(n=273/8.2%), those who had quit from occasional smoking, were consistently former smokers (quit before the baseline), or consistently never smokers as ’Non-current smokers’ (1,856/55.9%). The category ’Others’ included persons with miscellaneous smoking status (initiators, reducers, relapses, increasers, and other changes in smoking during the follow-up) (n=550/16.6%). No interpretation was made for the last group given its heterogeneity. The first three categories (persistent smokers, quitters, and non-current smoker) reflect decreasing exposure when moving from persistent smoking towards never smoking. The distribution of long-term smoking status is shown in Figure 1a.

#### Leisure-time physical activity

LTPA was calculated using the self-reported frequency (per month), duration (minutes per one session), and an average intensity in metabolic equivalents (METs). In addition, daily time of physically active commuting to work and back home was included^24^. From these measures, the total MET index of LTPA-h/week was calculated at both waves^14^.

### Covariates

At both waves, self-reported weight (to the nearest kilogram) and height (to the nearest centimeter) were used to calculate BMI (weight [kg]/ height [m]2). In addition to sex, age, and BMI, we included baseline socio-economic status, self-rated health, sleep problems, alcohol use (g/week), diet quality, psychological distress (General Health Questionnaire-12), and life satisfaction as covariates. The distributions of the covariates by long-term smoking status are described in Supplement Table 1.

### Analyses

#### Individual-based analyses

We report total numbers and percentages for categorical variables, means and standard deviations for continuous variables by long-term smoking status. Interactions between sex and smoking with WC change as well as between LTPA change and smoking with WC change were tested using Likelihood-ratio test comparing two nested models with and without interaction terms. Baseline age, sex, BMI, and WC were adjusted in the analyses. Interaction was considered if p<0.15. As there was no sex by smoking-interaction (p=0.375) men and women were pooled in the analyses. Because there was no interaction between LTPA and smoking status on WC change (p=0.33), all smoking status groups were pooled together when analyzing associations of LTPA with WC change, yet the analysis was adjusted for smoking status.

The effects of smoking and LTPA on WC change were estimated by modelling follow-up WC adjusted for baseline WC in linear regression models providing beta coefficients (β) and 95% Confidence Intervals (CI). Normality of WC distributions between the groups (sex and smoking status) were tested using the Shapiro-Wilk test of normality and equality of WC variances between the groups by the Levene’s robust test centered at the median.

To examine the association of smoking status (independent variable) with WC at follow-up (dependent variable), standard linear regression models (procedure reg in Stata) were performed as follows: model 1: adjusted for sex, age, and baseline WC; model 2: adjusted for sex, age, baseline WC, and baseline BMI; model 3: adjusted for all covariates (sex, age, baseline WC, baseline BMI, diet, baseline LTPA, life satisfaction, sleep problems, GHQ12, alcohol use, socioeconomic status, and self-rated health).

For estimating associations of LTPA (independent variable) with WC at follow-up (dependent variable) linear regression models were performed as follows: model 1: adjusted for sex, age, and baseline WC; model 2: adjusted for sex, age, baseline WC, and baseline BMI; model 3: adjusted for sex, age, baseline WC, baseline BMI, and smoking status.

As the primary sampling unit has been the twin pair, twins within a pair are not statistically independent observations. Therefore, standard errors in individual-based analyses were corrected using a simple robust variance estimator for cluster-correlated data by using a robust variance estimator (cluster option in Stata software)^25^.

#### Within-pair analyses

We conducted within-pair analyses (n=1,050 pairs) for the associations of smoking status (categorical independent variable) with WC (continuous dependent variable) and of LTPA (continuous independent variable) with WC (continuous dependent variable) using linear fixed-effects regression models (procedure xtreg, fe option). Twin pairs commonly share the same childhood environment and experiences. DZ pairs are genetically full siblings, whereas MZ pairs, are identical at their genomic sequence^26^. In the common use of fixed-effects models, estimates of exposure-outcome associations are derived from longitudinal variation in exposure within individuals, everyone being their own control. The models can also be used to conduct within-pair analyses of twin data, in which exposed twins are compared with their unexposed co-twins^27^. The twin pair design accounts for sex and age, as well as shared familial effects, whether measured or unmeasured in all twin pairs. In DZ pairs, there is residual genetic confounding, while genetic effects are fully controlled for in MZ pairs.

The results from within-pair analyses are informative when they are compared with the results from individual-based analyses^28^. If familial confounding plays a significant role, we should see an association among all individuals, but less or none within twin pairs. Examining MZ and DZ pairs separately informs whether familial confounding is due to shared genes or environment. Since DZ pairs can be of the same-sex (SS) or of opposite-sex (OS) (male-female), post-hoc analyses were conducted also for the SSDZ and OSDZ pairs separately.

Stata SE version 18 (StataCorp, College Station, Texas, USA) was used for all analyses. In the main analyses two-tailed p<0.05 was regarded as statistically significant.

## RESULTS

### Individual-based analyses

Among all 3,322 participants more than half (55,9%, n=1,856) were non-current smokers, 8.2% (n=273) were quitters from baseline daily smoking, 19.3% (n=643) were persistently smoking daily or occasionally, and 16.6% (n=550) were categorized as ‘others’ (Figure 1a, Supplement Table 2). During the follow-up, 36.4% of baseline daily smokers quit smoking.

The mean increase in WC during 10-year follow-up was on average 6.5 cm (SD 8.1), and it varied from 6.1 cm (SD 7.9) among non-current smokers to 8.4 cm (SD 8.1) among those who quit daily smoking during the follow-up. The detailed values for WC, weight, and BMI at baseline, at the end of follow-up, plus the change of them during the follow-up are described in the Supplement Table 2. Box-plots with the median for change in WC with 75^th^ percentiles by smoking status are illustrated in the Supplement Figure 2. Compared to those who continued smoking, WC increase was statistically significant only among those who quit daily smoking (multiple adjusted β 2.04; 95% CI 0.94, 3.14) (Table 1).

**Table 1.** Individual-based associations of long-term smoking status with waist circumference (cm) change during 10-year follow-up in the linear regression analyses* in the FinnTwin16 cohort (n=3,322).

| Long-term smoking status | Model 1<br>$\beta$ (95% CI) | Model 2<br>$\beta$ (95% CI) | Model 3<br>$\beta$ (95% CI) |
| --- | --- | --- | --- |
| Persistent smokers (n=643)<sup>a</sup> | Reference | Reference | Reference |
| Quitters (n=273)<sup>b</sup> | 1.74 (0.60, 2.87) | 1.76 (0.66, 2.87) | 2.04 (0.94, 3.14) |
| Non-current smokers (n=1,856)<sup>c</sup> | -0.52 (-1.25, 0.21) | -0.51 (-1.23, 0.21) | 0.08 (-0.67, 0.83) |

\*Linear Regression model including a robust variance estimator to adjust for the non-independence of observations within twin pairs. In all models, number of included individuals is 3,322. Smoking status 'other' (n=550) is in the model but not shown.
<sup>a</sup> Persistent daily or occasional smoking; <sup>b</sup> Quitting from daily smoking; <sup>c</sup> Quitting from occasional smoking, consistent former smoking, never smoking.
Model 1: adjusted for sex, age and baseline WC.
Model 2: adjusted for sex, age, baseline WC and baseline BMI.
Model 3: adjusted for sex, age, baseline WC, baseline BMI, diet, baseline LTPA, life satisfaction, sleep problems, GHQ12, alcohol use, socioeconomic status, and self-rated health.
$\beta$ = linear regression coefficients; CI = confidence intervals; BMI=body mass index; GHQ=General Health Questionnaire; LTPA= leisure-time physical activity; WC=waist circumference.

There was decrease in LTPA during the follow-up in all other smoking status groups, except among those who quit daily smoking, where the mean LTPA increase was 5.0 MET-h/week (SD 35.9). The detailed values for LTPA by smoking status are shown in the Supplement Table 2. Independently of smoking status, each additional MET-h/week was associated with less WC increase (adjusted β - 0.06; 95% CI -0.07, -0.05) (Table 2).

**Table 2.** Individual-based associations of waist circumference (cm) change during 10-year follow-up in relation to baseline leisure-time physical activity and change in leisure-time physical activity during the 10-year follow-up in the linear regression analyses* in the FinnTwin16 sample (n=3,322).

| **Leisure-time physical activity** | **Model 1**<br>$\beta$ (95% CI) | **Model 2**<br>$\beta$ (95% CI) | **Model 3**<br>$\beta$ (95% CI) |
| --- | --- | --- | --- |
| At baseline (MET-h/wk) | -0.04 (-0.05, -0.03) | -0.05 (-0.06, -0.04) | -0.05 (-0.06, -0.04) |
| Change in LTPA (MET-h/wk) | -0.06 (-0.07, -0.05) | -0.06 (-0.07, -0.05) | -0.06 (-0.07, -0.05) |

\*Linear Regression model including a robust variance estimator to adjust for the non-independence of observations within twin pairs. In all models, number of included individuals is 3,322.
Model 1: adjusted for sex, age and baseline WC.
Model 2: adjusted for sex, age, baseline WC and baseline BMI.
Model 3: adjusted for sex, age, baseline WC, baseline BMI, and smoking status.
$\beta$ = linear regression coefficients; CI = confidence intervals; BMI=body mass index; LTPA= leisure-time physical activity; MET-h/wk= metabolic equivalent hours per week; WC=waist circumference.

#### Within-pair analyses

There were 45 pairs (34 DZ pairs; 11 MZ pairs) where both co-twins were current smokers at baseline and one twin within a pair quit smoking during follow-up and the other one did not (Supplement Table 3). Among 11 MZ pairs, smoking cessation was not associated with WC increase (Table 3 and Supplement Figure 3). In other words, the co-twin who had quit smoking did not have increased WC if compared to his/her co-twin who continued daily smoking. Among 34 DZ pairs, the co-twin who quit smoking had a larger WC increase (Supplement Figure 3). Among the SSDZ pairs, there was no difference in WC change between the twin who quit smoking and the co-twin who did not (Table 3). A difference in WC change was found in the OSDZ twin pairs, but as sex is a major confounder that we cannot control in the within-pair analyses of OSDZ pairs, we do not know whether the effect is due to quitting or due to sex-associated factors that differ between the sister and brother in such pairs. The numbers of twin pairs concordant and discordant for smoking status are shown in Supplemental Table 3.

**Table 3.** Estimates for the association of smoking status with waist circumference (cm) change during 10-year follow-up within-pairs for 660 dizygotic pairs, and 390 monozygotic pairs.

| Long-term smoking status | Within DZ pairs<br>n=660 pairs<br>$\beta$ (95% CI) | Within OSDZ pairs<br>N=332 pairs<br>$\beta$ (95% CI) | Within SSDZ pairs<br>N=328 pairs<br>$\beta$ (95% CI) | Within MZ pairs<br>n=390 pairs<br>$\beta$ (95% CI) |
| --- | --- | --- | --- | --- |
| Persistent smokers | Reference | Reference | Reference | Reference |
| Quitters | 2.61 (0.28, 4.94) | 4.14 (0.93, 7.35) | 0.85 (-2.52, 4.21) | -2.64 (-5.88, 0.60) |
| Non-current smokers | -0.25 (-1.90, 1.41) | 0.09 (-2.11, 2.29) | -0.23 (-2.83, 2.37) | -2.10 (-3.96, -0.23) |

Linear regression fixed-effects beta coefficients ( $\beta$ ) with 95% confidence intervals (CI). Estimates adjusted for sex, baseline age, and baseline waist circumference.
DZ = dizygotic; MZ = monozygotic; OSDZ = opposite-sex dizygotic; SSDZ = same-sex dizygotic; WC=waist circumference.

Concerning LTPA in relation to WC, within-pair analyses replicated the finding of the individual-based analysis suggesting that each additional MET-h/week of LTPA decreases the risk for WC increase. This association remained similar and significant both within DZ pairs (β=-0.04 (-0.06, -0.02) as well as within MZ pairs (β=-0.05; -0.07, -0.02) (Table 4). The effect sizes are very similar in the analysis of all individuals (Table 2) and within-pairs (Table 4).

**Table 4.** Adjusted estimates for the association of leisure-time physical activity change with waist circumference (cm) change during 10-year follow-up within 660 dizygotic pairs, and 390 monozygotic pairs.

| Leisure-time physical activity | Within DZ pairs<br>n=660 pairs<br>$\beta$ (95% CI) | Within MZ pairs<br>n=390 pairs<br>$\beta$ (95% CI) |
| --- | --- | --- |
| Change in LTPA (MET-h/wk) | -0.04 (-0.06, -0.02) | -0.05 (-0.07, -0.02) |

Linear regression fixed-effects beta coefficients ( $\beta$ ) with 95% confidence intervals. Estimates adjusted for age at baseline, sex, baseline waist circumference, baseline body mass index, and leisure-time physical activity at baseline.
LTPA= leisure-time physical activity; WC=waist circumference; DZ = dizygotic; MZ = monozygotic; CI = confidence intervals; MET-h/wk= metabolic equivalent hours per week.

## DISCUSSION

In summary, this study showed a decrease in the prevalence of smoking, reduced amount of LTPA and increasing trend in abdominal obesity among the adult twin cohort at the end of the 10-year follow-up. According to multiple adjusted individual-based analyses, quitting daily smoking was robustly associated with an increase in WC when compared to those who continued smoking.

However, the within-pair analysis led to more ambiguous estimates. This may reflect shared familial confounding and/or insufficient statistical power due to small sample of smoking discordant twins in the within-pair analysis^27^. Further, LTPA appears to function as a protective factor on post-cessation WC increase. Namely, when comparing the estimates from the individual-based and within-pair analyses among all, and especially, among genetically identical twin pairs, the estimates remained stable. This illustrates power of twin design to strengthen the evidence for the causal nature of an association when there is enough statistical power in the data.

In this cohort, about every third baseline daily smoker quit during the follow-up. Quitting smoking in our data follows the global trend of smoking prevalence in high income western countries^29^. In our data, both BMI and WC increased during the 10-year follow-up, although the individual variability was wide. Our results support previous findings about weight and BMI development in different smoking groups, including post-cessation weight development^30^. In our cohort, the mean increase among those who quit daily smoking was 8.4 cm during the 10 years. Although longitudinal studies about the effect of smoking status on WC are rare, there is evidence that on average, WC increases 3.9 cm during the first post-cessation year^11^. In a Danish study^11^, 40% of quitters and 15% of those continuing smoking increased their WC at least 5 cm during one-year follow-up. In our cohort, when we compared WC change among those quitting daily smoking (8.4 cm) to those who continued smoking (6.6 cm), the WC increase was about 2 cm more in quitters.

Several mechanisms for post-cessation weight gain or waist increase have been proposed. First, that nicotine increases energy expenditure about 10%, especially during exercise and after eating^31^. Second, that smokers have less appetite and thus, individuals who quit smoking tend to increase their body weight as their appetite improves^1–4^ ^6^. In terms of health consequences, even 2 cm increase in WC increases risk for insulin resistance and cardiometabolic diseases^1^ ^13^ ^32^.

However, there is growing evidence suggesting that continuing smoking also increases the risk for abdominal obesity.^13^ ^30^ ^33^ Interestingly, there is support for a notion that nicotine itself may lead to insulin resistance, hence increasing the risk of type 2 diabetes^1^ ^34^. Therefore, the notion of smoking as ‘weight control method’ appears to be misleading.

In our twin sample, quitting daily smoking was robustly associated with WC increase when several potential confounders were controlled for. However, when familial factors were controlled for, the association did not remain robust. In other words, our twin data did not provide evidence to support a causal association between smoking cessation and risk for abdominal obesity. There are several factors that may affect such as lack of statistical power and confounding by shared genetic and/or environmental factors. Concerning statistical power, number of smoking discordant twin pairs was low. Especially, there were only 45 pairs (11 MZ pairs) where one twin was persistent smoker while his/her co-twin was quitter. Therefore, lack of statistical power does not allow to make strong conclusions. However, it is also not possible to reject familial confounding, especially, because the point estimates of individual level were not replicated among twin pairs. Genetic factors confounding the association between smoking cessation and weight gain has also been found in adult male twins^35^. Smoking was associated with lower BMI, and smoking cessation with higher BMI in the multicenter study of 150 000 twins^36^. However, the ‘net effect’ of smoking cessation on weight was not more than an average of 0.7 kg/m^2^. This evidence supports previous suggestions that after smoking cessation, individual’s weight returns in long run to the same weight-age trajectory as observed in never smokers^31^ ^36^.

The mean amount of LTPA decreased in all smoking status groups except among quitters from daily smoking. Our results, in general, are in line with previous findings demonstrating a decrease in LTPA by age^37^. The association between smoking status and WC increase is moderated by sedentary behavior so that current and former smokers spend more sedentary time compared with never smokers. Sedentary ever smokers have also highest BMI and WC values^12^. Furthermore, heavy smoking among sedentary population increases BMI, that in turn further decreases LTPA level^38^.

In our individual-based analysis, independently of smoking status, every increase in LTPA MET- h/week decreased the likelihood for WC increase by 0.06 cm. The effect is small since to prevent one cm WC increase, a person should have performed at least 20 MET-h additional exercise per week (equal to 4 hours walking at speed of 5 km/h). From clinical perspective, even this small effect can be important when we take into account the positive association of physical activity with other lifestyle factors as sleep^30^ affecting also WC^39^. Physical activity has, in any case, a role in preventing and treatment of obesity^40^. Importantly, in our within-pair analyses, the effect sizes of the association remained consistent. To summarize, our results suggest that the association of LTPA with WC change may be independent of familial predisposition as the effect sizes in the within-pair analyses of MZ pairs were consistent with the effect sizes for individuals.

Exercise can prevent weight gain in general^14^ and after smoking cessation^15^ but less has been known about the impact of LTPA on abdominal obesity and related outcomes. In a recent Finnish twin study including long-term LTPA discordant MZ pairs, the more active co-twins had decreased amount of body fat, visceral fat, and liver fat, plus more advantageous health-related benefits compared to their less physically active co-twins^41^. Previous evidence about the role of physical exercise in preventing post-cessation WC increase is limited^15^. In a smoking cessation trial including nicotine replacement therapy, counselling, and exercise, abstinence was followed by moderate weight gain, but not by increase in visceral fat^16^.

Earlier studies have reported that smoking cessation is associated with post-cessation-related obesity which in turn might contribute to worsening in lipid parameters and insulin resistance^1^ ^32, 42^. If this association is causal, it is essential to improve weight management interventions during smoking cessation. This is especially important for people living with diabetes^42^. While there is no constant evidence that physical exercise during the quitting process might increase the successfulness of smoking cessation^43^, some physical exercise interventions have been associated with less weight gain during 12-month post-cessation follow-up^15^. Post-cessation weight management is also important in supporting smoking abstinence and in relapse prevention^44^. Whether longer behavioral and weight management interventions benefit successful cessation and reduce post-cessation abdominal obesity remains to be investigated^31^.

This study has several strengths. First, our study is based on longitudinal population-based data including a large sample of men and women. Second, we included several long-term smoking status groups. Third, WC was measured twice for estimating abdominal obesity. Fourth, the data allowed adjustment for several potential confounders. Finally, we utilized twin data providing a powerful design for testing familial confounding.

The main limitation is using self-reported data. We acknowledge that the self-measured WC can be less reliable to WC measured by study investigators following standard operating procedures. For example, self-measurement maybe less accurate depending on the chosen technique because the arms must remain low along the trunk. However, in general, self-reported values of weight, height, and WC have been shown relatively accurate for large cohort studies^23^. Further, smoking cessation was not biochemically verified, but the smoking questions were part of a larger survey. Thus, social desirability to indicate having quit may be less strong when using such a general health and well-being survey. We do not know how many times participants had changed their smoking behavior and when exactly they had quit smoking during the follow-up. However, there is evidence that most of the weight related changes usually occur during the first post-cessation years^3^ ^4^. Furthermore, the LTPA data did not classify the intensity of physical activity (sedentary, light, moderate, vigorous), which are common terms used to recommend types of physical activity in weight management services and in exercise prescription. We acknowledge this as a further study limitation.

## CONCLUSIONS

Smoking cessation seems to be associated with moderate increase in waist circumference, although familial influences may be involved in this process. Leisure-time physical activity appears to prevent waist circumference increase independently of familial influences. This finding highlights the importance of incorporating regular physical activity into lifestyle interventions, such as smoking cessation programs, where prevention of post-cessation risk for abdominal obesity may be beneficial.

## ETHICS APPROVAL

FinnTwin16 study followed the ethical standards and the Declaration of Helsinki. The Ethics Committees of the Hospital District of Helsinki and Uusimaa (113/E3/2001) and the Institutional Review Board of Indiana University, Bloomington, IN, USA approved the study. The Ethics Committee of the Central Finland Hospital district accepted the fifth data collection (5E/2010). At all waves of the study, twins gave their informed consent by returning the questionnaire.

## Data Availability

All data produced in the present study are available upon reasonable request to the authors.

## ACKNOWLEDGMENTS

We acknowledge all twin participants for their valuable co-operation along the years. This study was financially supported by the grants from the Juho Vainio Foundation (to Korhonen) and from the Sigrid Juselius Foundation and Research Council of Finland (grant # 352792 to Kaprio).

## AUTHOR CONTRIBUTIONS

Maarit Piirtola: Conceptualization; Data curation; Formal analysis; Investigation; Methodology; Software; Validation; Visualization; Writing - original draft and review & editing.

Eeva-Liisa Filippone: Conceptualization, Writing - review & editing. Anu Ranjit: Conceptualization, Writing - review & editing.

Taru Kinnunen: Conceptualization, Writing - review & editing.

Jaakko Kaprio: Conceptualization; Data curation; Funding acquisition; Investigation; Methodology; Project administration; Resources; Supervision; Writing - review & editing.

Tellervo Korhonen: Conceptualization; Data curation; Funding acquisition; Investigation; Methodology; Project administration; Resources; Supervision; Validation; Writing - review & editing.

## COMPETING INTEREST STATEMENT

The authors confirm that there are no competing interests to be disclosed.

## DATA AVAILABILITY STATEMENT

Finnish Twin Cohort data is available through the Institute for Molecular Medicine Finland (FIMM) Data Access Committee (DAC) for authorized researchers who have IRB/ethics approval and an institutionally approved study plan. To ensure the protection of privacy and compliance with national data protection legislation, a data use/transfer agreement is needed, the content and specific clauses of which will depend on the nature of the requested data.

## COMPETING INTEREST STATEMENT

None disclosed

## FUNDING

This study was financially supported by the grants from the Juho Vainio Foundation (to Korhonen) and from the Sigrid Juselius Foundation and Research Council of Finland (grant # 352792 to Kaprio).

## Notes

### Competing Interest Statement

The authors have declared no competing interest.

### Funding Statement

This study was financially supported by the grants from the Juho Vainio Foundation (Tellervo Korhonen) and from the Sigrid Juselius Foundation and Research Council of Finland (grant # 352792 to Jaakko Kaprio).

### Author Declarations

FinnTwin16 study followed the accepted ethical standards and the Declaration of Helsinki. The Ethics Committees of the Hospital District of Helsinki and Uusimaa and the Institutional Review Board of Indiana University, Bloomington, IN, USA approved the FinnTwin16 study. The Ethics Committee of the Central Finland Hospital district accepted the fifth wave of data collection. At all waves of the FinnTwin16 study, twins gave their in-formed consent by returning the questionnaire.

### Summary of Updates

Statistical analyses have been updated throughout the paper. Results and conclusions have been revised accordingly. All tables and figures have been updated.

## REFERENCES

1. Harris KK, Zopey M, Friedman TC. Metabolic effects of smoking cessation. Nat Rev Endocrinol 2016;12(5):299–308. doi: 10.1038/nrendo.2016.32 [published Online First: 20160304]

2. Tian J, Venn A, Otahal P, et al. The association between quitting smoking and weight gain: a systematic review and meta-analysis of prospective cohort studies. Obesity reviews : an official journal of the International Association for the Study of Obesity 2015;16(10):883–901. doi: 10.1111/obr.12304 [published Online First: 20150626]

3. Aubin HJ, Farley A, Lycett D, et al. Weight gain in smokers after quitting cigarettes: meta-analysis. BMJ (Clinical research ed) 2012;345:e4439. doi: 10.1136/bmj.e4439 [published Online First: 20120710]

4. Spring B, Howe D, Berendsen M, et al. Behavioral intervention to promote smoking cessation and prevent weight gain: a systematic review and meta-analysis. Addiction 2009;104(9):1472–86. doi: 10.1111/j.1360-0443.2009.02610.x [published Online First: 20090622]

5. Locatelli I, Collet TH, Clair C, et al. The joint influence of gender and amount of smoking on weight gain one year after smoking cessation. Int J Environ Res Public Health 2014;11(8):8443–55. doi: 10.3390/ijerph110808443 [published Online First: 20140818]

6. Veldheer S, Yingst J, Zhu J, et al. Ten-year weight gain in smokers who quit, smokers who continued smoking and never smokers in the United States, NHANES 2003-2012. *International journal of obesity* 2015;39(12):1727-32. doi: 10.1038/ijo.2015.127 [published Online First: 20150709]

7. Siahpush M, Singh GK, Tibbits M, et al. It is better to be a fat ex-smoker than a thin smoker: findings from the 1997-2004 National Health Interview Survey-National Death Index linkage study. Tobacco control 2014;23(5):395–402. doi: 10.1136/tobaccocontrol-2012-050912 [published Online First: 20130410]

8. Hu Y, Zong G, Liu G, et al. Smoking Cessation, Weight Change, Type 2 Diabetes, and Mortality. The New England journal of medicine 2018;379(7):623–32. doi: 10.1056/NEJMoa1803626

9. Reis JP, Allen N, Gunderson EP, et al. Excess body mass index- and waist circumference-years and incident cardiovascular disease: the CARDIA study. Obesity 2015;23(4):879–85. doi: 10.1002/oby.21023 [published Online First: 20150309]

10. Cerhan JR, Moore SC, Jacobs EJ, et al. A pooled analysis of waist circumference and mortality in 650,000 adults. Mayo Clinic proceedings 2014;89(3):335–45. doi: 10.1016/j.mayocp.2013.11.011

11. Pisinger C, Jorgensen T. Waist circumference and weight following smoking cessation in a general population: the Inter99 study. Preventive medicine 2007;44(4):290–5. doi: 10.1016/j.ypmed.2006.11.015 [published Online First: 20070111]

12. Kaufman A, Augustson EM, Patrick H. Unraveling the Relationship between Smoking and Weight: The Role of Sedentary Behavior. Journal of obesity 2012;2012:735465. doi: 10.1155/2012/735465 [published Online First: 20110927]

13. Terry JG, Hartley KG, Steffen LM, et al. Association of smoking with abdominal adipose deposition and muscle composition in Coronary Artery Risk Development in Young Adults (CARDIA) participants at mid-life: A population-based cohort study. PLoS medicine 2020;17(7):e1003223. doi: 10.1371/journal.pmed.1003223 [published Online First: 20200721]

14. Piirtola M, Kaprio J, Waller K, et al. Leisure-time physical inactivity and association with body mass index: a Finnish Twin Study with a 35-year follow-up. International journal of epidemiology 2017;46(1):116–27. doi: 10.1093/ije/dyw007

15. Hartmann-Boyce J, Theodoulou A, Farley A, et al. Interventions for preventing weight gain after smoking cessation. The Cochrane database of systematic reviews 2021;10(10):CD006219. doi: 10.1002/14651858.CD006219.pub4 [published Online First: 20211006]

16. Prapavessis H, De Jesus S, Fitzgeorge L, et al. Anthropometric and body composition changes in smokers vs abstainers following an exercise-aided pharmacotherapy smoking cessation trial for women. Addictive behaviors 2018;85:125–30. doi: 10.1016/j.addbeh.2018.06.003 [published Online First: 20180607]

17. Tynkkynen NP, Tormakangas T, Palviainen T, et al. Associations of polygenic inheritance of physical activity with aerobic fitness, cardiometabolic risk factors and diseases: the HUNT study. European journal of epidemiology 2023;38(9):995–1008. doi: 10.1007/s10654-023-01029-w [published Online First: 20230821]

18. Zadro JR, Shirley D, Andrade TB, et al. The Beneficial Effects of Physical Activity: Is It Down to Your Genes? A Systematic Review and Meta-Analysis of Twin and Family Studies. Sports Med Open 2017;3(1):4. doi: 10.1186/s40798-016-0073-9 [published Online First: 20170110]

19. Sillanpaa E, Palviainen T, Ripatti S, et al. Polygenic Score for Physical Activity Is Associated with Multiple Common Diseases. Medicine and science in sports and exercise 2022;54(2):280–87. doi: 10.1249/MSS.0000000000002788

20. Boomsma D, Busjahn A, Peltonen L. Classical twin studies and beyond. Nature reviews Genetics 2002;3(11):872-82. doi: 10.1038/nrg932 [published Online First: 2002/11/05]

21. Kaidesoja M, Aaltonen S, Bogl LH, et al. FinnTwin16: A Longitudinal Study from Age 16 of a Population-Based Finnish Twin Cohort. Twin research and human genetics : the official journal of the International Society for Twin Studies 2019;22(6):530–39. doi: 10.1017/thg.2019.106 [published Online First: 20191203]

22. Sarna S, Kaprio J, Sistonen P, et al. Diagnosis of twin zygosity by mailed questionnaire. Human heredity 1978;28(4):241–54. doi: 10.1159/000152964

23. Tuomela J, Kaprio J, Sipila PN, et al. Accuracy of self-reported anthropometric measures - Findings from the Finnish Twin Study. Obes Res Clin Pract 2019;13(6):522–28. doi: 10.1016/j.orcp.2019.10.006 [published Online First: 20191121]

24. Raza W, Krachler B, Forsberg B, et al. Health benefits of leisure time and commuting physical activity: A meta-analysis of effects on morbidity. Journal of Transport & Health 2020;18(September):100873. doi: 10.1016/j.jth.2020.100873

25. Williams RL. A note on robust variance estimation for cluster-correlated data. Biometrics 2000;56(2):645–6. doi: 10.1111/j.0006-341x.2000.00645.x [published Online First: 2000/07/06]

26. Jonsson H, Magnusdottir E, Eggertsson HP, et al. Differences between germline genomes of monozygotic twins. Nature genetics 2021;53(1):27–34. doi: 10.1038/s41588-020-00755-1 [published Online First: 20210107]

27. Gustavson K, Torvik FA, Davey Smith G, et al. Familial confounding or measurement error? How to interpret findings from sibling and co-twin control studies. European journal of epidemiology 2024;39(6):587–603. doi: 10.1007/s10654-024-01132-6 [published Online First: 20240616]

28. McGue M, Osler M, Christensen K. Causal Inference and Observational Research: The Utility of Twins. Perspectives on psychological science : a journal of the Association for Psychological Science 2010;5(5):546–56. doi: 10.1177/1745691610383511

29. Collaborators GBDT. Spatial, temporal, and demographic patterns in prevalence of smoking tobacco use and attributable disease burden in 204 countries and territories, 1990-2019: a systematic analysis from the Global Burden of Disease Study 2019. Lancet 2021;397(10292):2337-60. doi: 10.1016/S0140-6736(21)01169-7 [published Online First: 20210527]

30. Behl TA, Stamford BA, Moffatt RJ. The Effects of Smoking on the Diagnostic Characteristics of Metabolic Syndrome: A Review. Am J Lifestyle Med 2023;17(3):397–412. doi: 10.1177/15598276221111046 [published Online First: 20220628]

31. Audrain-McGovern J, Benowitz NL. Cigarette smoking, nicotine, and body weight. Clinical pharmacology and therapeutics 2011;90(1):164–8. doi: 10.1038/clpt.2011.105 [published Online First: 20110601]

32. Driva S, Korkontzelou A, Tonstad S, et al. The Effect of Smoking Cessation on Body Weight and Other Metabolic Parameters with Focus on People with Type 2 Diabetes Mellitus. Int J Environ Res Public Health 2022;19(20) doi: 10.3390/ijerph192013222 [published Online First: 20221014]

33. Carrasquilla GD, Garcia-Urena M, Romero-Lado MJ, et al. Estimating causality between smoking and abdominal obesity by Mendelian randomization. Addiction 2024;119(6):1024–34. doi: 10.1111/add.16454 [published Online First: 20240320]

34. Edstorp J, Ahlqvist E, Alfredsson L, et al. Incidence of LADA and Type 2 Diabetes in Relation to Tobacco Use and Genetic Susceptibility to Type 2 Diabetes and Related Traits: Findings From a Swedish Case-Control Study and the Norwegian HUNT Study. Diabetes Care 2023;46(5):1028–36. doi: 10.2337/dc22-2284

35. Swan GE, Carmelli D. Characteristics associated with excessive weight gain after smoking cessation in men. American journal of public health 1995;85(1):73–7. doi: 10.2105/ajph.85.1.73 [published Online First: 1995/01/01]

36. Piirtola M, Jelenkovic A, Latvala A, et al. Association of current and former smoking with body mass index: A study of smoking discordant twin pairs from 21 twin cohorts. PloS one 2018;13(7):e0200140. doi: 10.1371/journal.pone.0200140 [published Online First: 20180712]

37. Guthold R, Stevens GA, Riley LM, et al. Worldwide trends in insufficient physical activity from 2001 to 2016: a pooled analysis of 358 population-based surveys with 1.9 million participants. Lancet Glob Health 2018;6(10):e1077–e86. doi: 10.1016/S2214-109X(18)30357-7 [published Online First: 20180904]

38. Dvorak RD, Del Gaizo AL, Engdahl RM, et al. Tobacco use and body mass index: mediated effects through physical inactivity. Journal of health psychology 2009;14(7):919–23. doi: 10.1177/1359105309341005 [published Online First: 2009/09/30]

39. Sperry SD, Scully ID, Gramzow RH, et al. Sleep Duration and Waist Circumference in Adults: A Meta-Analysis. Sleep 2015;38(8):1269–76. doi: 10.5665/sleep.4906 [published Online First: 20150801]

40. Cornier MA. A review of current guidelines for the treatment of obesity. Am J Manag Care 2022;28(15 Suppl):S288-S96. doi: 10.37765/ajmc.2022.89292

41. Kujala UM, Leskinen T, Rottensteiner M, et al. Physical activity and health: Findings from Finnish monozygotic twin pairs discordant for physical activity. Scandinavian journal of medicine & science in sports 2022;32(9):1316–23. doi: 10.1111/sms.14205 [published Online First: 20220707]

42. Walicka M, Russo C, Baxter M, et al. Impact of stopping smoking on metabolic parameters in diabetes mellitus: A scoping review. World J Diabetes 2022;13(6):422–33. doi: 10.4239/wjd.v13.i6.422

43. Ussher MH, Taylor AH, Faulkner GE. Exercise interventions for smoking cessation. The Cochrane database of systematic reviews 2014;8:Cd002295. doi: 10.1002/14651858.CD002295.pub5 [published Online First: 2014/08/30]

44. Hankey C, Leslie W. Obesity: is weight gain after smoking cessation an important concern? Nat Rev Endocrinol 2012;8(11):630–2. doi: 10.1038/nrendo.2012.175 [published Online First: 20121002]

