## Supplemental Tables and Figures for "Associations of Smoking Status and Leisure-time Physical Activity with Waist Circumference Change – Ten-year Follow-up among Twin Adults"

**Supplement Table 1.** Baseline distributions of covariates among individuals (n=3,322) by their long-term smoking status.

| Long-term smoking status | Persistent smokers [a] n=643 (19.3%) | Quitters [b] n=273 (8.2%) | Non-current smokers [c] n=1,856 (55.9%) | Others [d] n=550 (16.6%) |
|---|---|---|---|---|
| **Categorical variables** | **n (%)** | **n (%)** | **n (%)** | **n (%)** |
| **Sex** | | | | |
| Men (n=1,527) | 333 (51.8) | 141 (51.6) | 781 (42.1) | 272 (49.5) |
| Women (n=1,795) | 310 (48.2) | 132 (48.4) | 1,075 (57.9) | 278 (50.5) |
| **Socio-economic status** | | | | |
| Very good/fairly good | 180 (28.0) | 96 (35.2) | 665 (35.8) | 186 (33.8) |
| Average | 278 (43.2) | 120 (44.0) | 812 (43.8) | 229 (41.6) |
| Fairly bad/very bad | 185 (28.8) | 57 (20.9) | 379 (20.4) | 135 (24.5) |
| **Self-rated health** | | | | |
| Excellent /very good | 488 (75.9) | 224 (82.1) | 1,634 (88.0) | 477 (86.7) |
| Good | 136 (21.2) | 46 (16.8) | 190 (10.2) | 65 (11.8%) |
| Rather poor/very poor | 19 (3.0) | 3 (1.1) | 32 (1.7) | 8 (1.5) |
| **Sleep problems** | | | | |
| Not at all/seldom | 441 (68.6) | 201 (73.6) | 1,405 (75.7) | 414 (75.3) |
| Once a week/more often | 202 (31.4) | 72 (26.4) | 451 (24.3) | 136 (24.7) |
| **Continuous variables** | **mean (SD)** | **mean (SD)** | **mean (SD)** | **mean (SD)** |
| **Age** | 24.5 (0.9) | 24.5 (1.0) | 24.4 (0.9) | 24.4 (0.9) |
| **Alcohol use** (g/week) | 96.1 (121.6) | 78.0 (88.5) | 43.1 (60.3) | 68.4 (87.1) |
| **Diet quality** (DQS, sum score) | 7.0 (2.1) | 7.7 (2.1) | 8.2 (2.1) | 7.7 (2.2) |
| **Psychological distress** (GHQ12, sum score) | 11.3 (5.5) | 11.0 (5.7) | 10.4 (5.2) | 10.3 (4.7) |
| **Life satisfaction** (sum score) | 8.9 (3.2) | 9.0 (3.4) | 8.3 (2.9) | 8.3 (3.0) |

[a] Persistent daily or occasional smoking; [b] Quitting from daily smoking; [c] Quitting from occasional smoking, consistent former smoking, never smoking; [d] Miscellaneous smoking status (initiators, reducers, relapses, increases and other changes in smoking status during the follow-up). DQS=Diet quality score; GHQ12= General Health Questionnaire 12.

**Supplement Table 2.** Means (SD) of anthropometric measures and leisure-time physical activity at baseline and follow-up by smoking status among individuals (n=3,322).

| **Long-term smoking status** | **Persistent smokers** [a] | **Quitters** [b] | **Non-current smokers** [c] | **Others** [d] |
|---|---|---|---|---|
| | n=643 | n=273 | n=1,856 | n=550 |
| **Measure** | | | | |
| Height (cm) baseline | 172.4 (9.0) | 172.4 (9.3) | 171.9 (9.2) | 172.9 (9.1) |
| Height (cm) follow-up | 172.4 (9.1) | 172.4 (9.2) | 171.9 (9.2) | 172.9 (9.2) |
| Weight (kg) baseline | 69.2 (13.7) | 68.1 (12.7) | 67.8 (13.4) | 68.7 (13.9) |
| Weight (kg) follow-up | 74.7 (15.5) | 75.4 (15.5) | 72.9 (15.6) | 74.2 (15.9) |
| BMI ($kg/m^2$) baseline | 23.1 (3.5) | 22.8 (3.2) | 22.8 (3.3) | 22.8 (3.3) |
| BMI ($kg/m^2$) follow-up | 25.0 (4.1) | 25.3 (4.1) | 24.5 (4.3) | 24.7 (4.3) |
| WC (cm) baseline | 80.4 (10.9) | 79.4 (10.0) | 78.9 (10.3) | 79.5 (10.3) |
| WC (cm) follow-up | 87.0 (13.1) | 87.8 (12.3) | 84.9 (12.2) | 86.4 (12.8) |
| Δ WC (cm) | 6.6 (8.3) | 8.4 (8.1) | 6.1 (7.9) | 6.9 (8.4) |
| LTPA (MET-h/wk) baseline | 22.0 (28.9) | 20.4 (24.2) | 34.3 (35.5) | 31.0 (33.5) |
| LTPA (MET-h/wk) follow-up | 19.3 (25.3) | 25.4 (28.4) | 27.5 (29.1) | 24.7 (27.6) |
| Δ LTPA (MET-h/wk) | -3.2 (31.8) | 5.0 (35.0) | -8.2 (35.5) | -6.0 (35.3) |

[a] Persistent daily or occasional smoking.

[b] Quitting from daily smoking.

[c] Quitting from occasional smoking, consistent former smoking, never smoking.

[d] Miscellaneous smoking status (initiators, reducers, relapses, increases and other changes in smoking status during the follow-up).

**Supplement Table 3.** Distribution of numbers of all 1,050 pairs and of 390 monozygotic pairs by their pairwise smoking status. The smoking status of the first twin within a pair is represented by the table rows and the smoking status of the second twin within a pair by table columns. Concordant pairs are on the diagonal and discordant pairs are off diagonal.

| Smoking status | Persistent | Quitters | Non-current | Other |
|---|---|---|---|---|
| **All pairs** | | | | |
| Persistent smokers | 76 | 45 | 129 | 66 |
| Quitters | | 16 | 59 | 23 |
| Non-current smokers | | | 417 | 178 |
| Other | | | | 41 |
| **Monozygotic pairs** | | | | |
| Persistent smokers | 40 | 11 | 31 | 31 |
| Quitters | | 6 | 19 | 7 |
| Non-current smokers | | | 166 | 60 |
| Other | | | | 19 |

**PICTORIAL INSTRUCTION FOR WAIST CIRCUMFERENCE MEASUREMENT**

Finally, we ask you to measure round your waist with the measuring tape we have sent you earlier along with the invitation letter. Please stand up straight when measuring. Measure the slimmest point of your waist. If you have difficulties in finding it, measure as shown at the picture, the circumference situated in the middle of the lowest part of the ribs (A) and the upper part of the hip bone (B).

My waist measurement is _______ cm

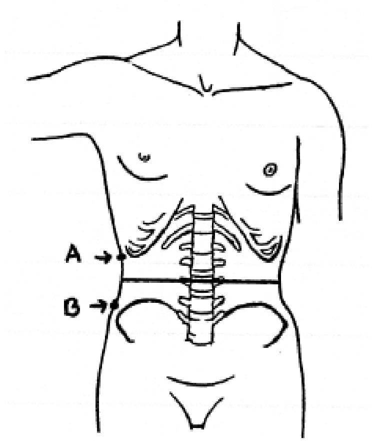


**Supplement Figure 1.** Pictorial instruction for waist circumference self-measurement.

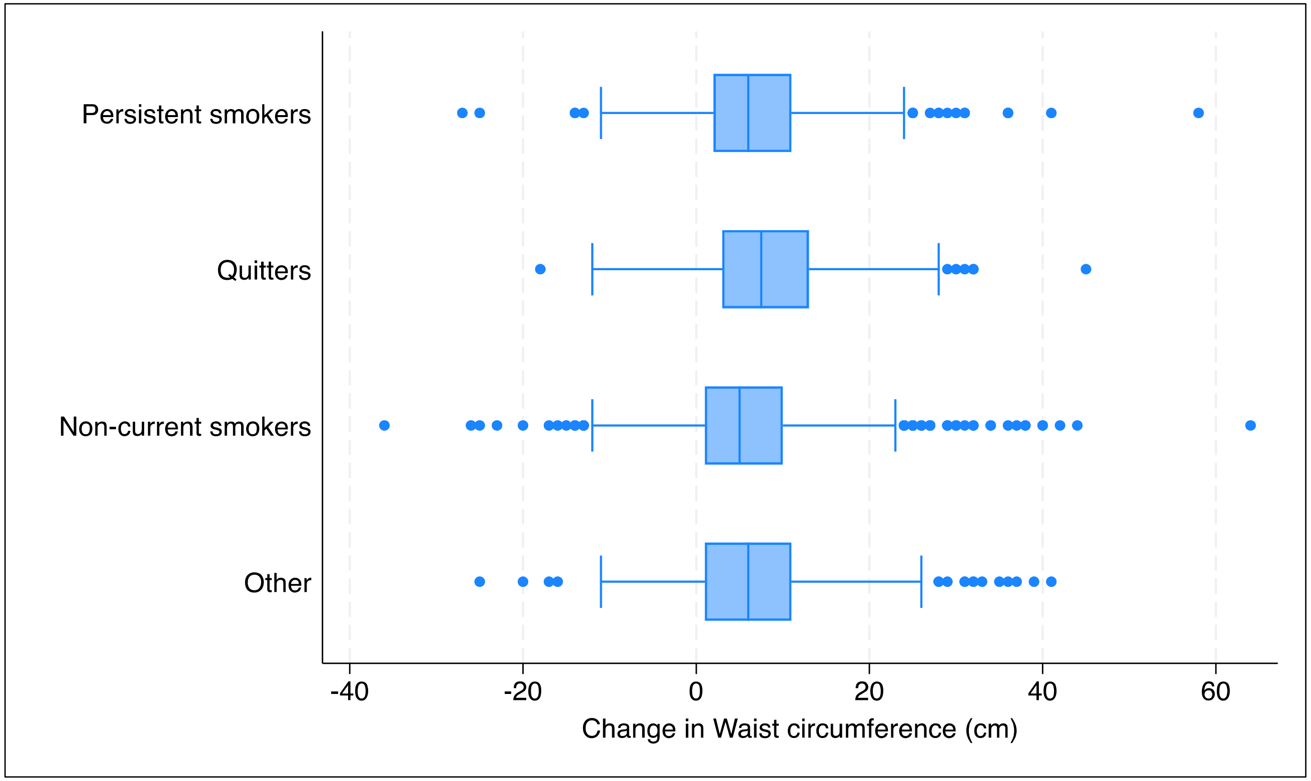


**Supplement Figure 2**. Box-plots with the median for change in waist circumference (WC) with 75$^{th}$ percentiles by smoking status during 10-year follow-up.

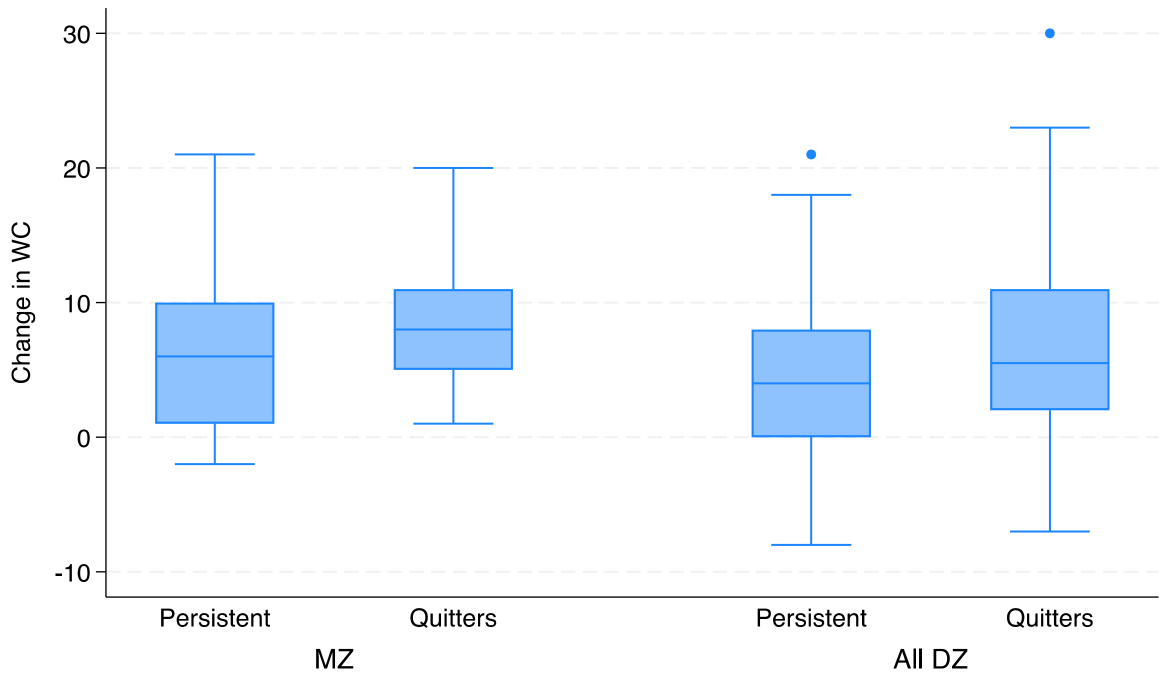

Change in WC
30
20
10
0
-10
Persistent
Quitters
Persistent
Quitters
MZ
All DZ

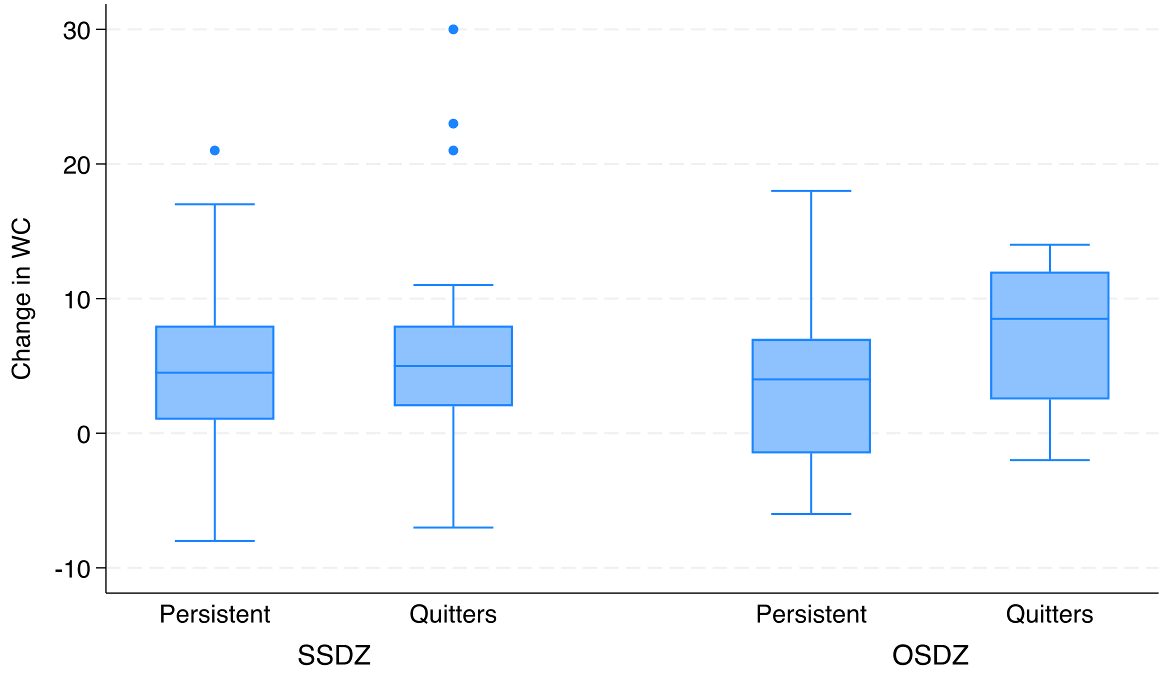


**Supplement Figure 3.** Change in waist circumference (WC) among 45 discordant twin pairs (persistent smokers *versus* quitters).

MZ = monozygotic (n=11 pairs); ALL DZ = All dizygotic (n=34 pairs); SSDZ=same-sex dizygotic (n=22 pairs); OSDZ=opposite-sex dizygotic (n=12 pairs)
